# Evaluation of ChatGPT’s Usefulness and Accuracy in Diagnostic Surgical Pathology

**DOI:** 10.1101/2024.03.12.24304153

**Authors:** Vincenzo Guastafierro, Devin Nicole Corbitt, Alessandra Bressan, Bethania Fernandes, Ömer Mintemur, Francesca Magnoli, Susanna Ronchi, Stefano La Rosa, Silvia Uccella, Salvatore Lorenzo Renne

**Affiliations:** Department of Biomedical Sciences, Humanitas University, Via Rita Levi Montalcini 4, 20090 Pieve Emanuele, Milan, Italy; IRCCS Humanitas Research Hospital, via Manzoni 56, 20089 Rozzano, Milan, Italy; Unit of Pathology, Department of Oncology, ASST Sette Laghi, Varese, Italy; Unit of Pathology, Department of Medicine and Technological Innovation, University of Insubria, Varese, Italy

## Abstract

ChatGPT is an artificial intelligence capable of processing and generating human-like language. ChatGPT’s role within clinical patient care and medical education has been explored; however, assessment of its potential in supporting histopathological diagnosis is lacking. In this study, we assessed ChatGPT’s reliability in addressing pathology-related diagnostic questions across 10 subspecialties, as well as its ability to provide scientific references. We created five clinico-pathological scenarios for each subspecialty, posed to ChatGPT as open-ended or multiple-choice questions. Each question either asked for scientific references or not. Outputs were assessed by six pathologists according to: 1) usefulness in supporting the diagnosis and 2) absolute number of errors. All references were manually verified. We used directed acyclic graphs and structural causal models to determine the effect of each scenario type, field, question modality and pathologist evaluation. Overall, we yielded 894 evaluations. ChatGPT provided useful answers in 62.2% of cases. 32.1% of outputs contained no errors, while the remaining contained at least one error (maximum 18). ChatGPT provided 214 bibliographic references: 70.1% were correct, 12.1% were inaccurate and 17.8% did not correspond to a publication. Scenario variability had the greatest impact on ratings, followed by prompting strategy. Finally, latent knowledge across the fields showed minimal variation. In conclusion, ChatGPT provided useful responses in one-third of cases, but the number of errors and variability highlight that it is not yet adequate for everyday diagnostic practice and should be used with discretion as a support tool. The lack of thoroughness in providing references also suggests caution should be employed even when used as a self-learning tool. It is essential to recognize the irreplaceable role of human experts in synthesizing images, clinical data and experience for the intricate task of histopathological diagnosis.

## Introduction - Background and significance

Nowadays, the global healthcare system is witnessing a paradoxical phenomenon whereby the incidence of neoplastic pathology presents a steadily increasing trend against a declining trend in the number of new pathologists entering the discipline, posing a unique challenge for pathology services worldwide.^1,2^ The development of high-resolution scanning devices has made it possible to digitalize conventional glass slides to produce digital slides – a process referred to as whole-slide imaging (WSI) – opening the door to the era of digital pathology.^3,4^ In turn, digital pathology has made it possible to streamline the workflow of pathologists, to make remote collaboration between colleagues easier, and to reduce sign-out time, all of which help to improve the performance of anatomic pathology services.^5^ The shift to digital slides, along with the availability of large datasets, has enabled the development and integration of artificial intelligence (AI) models, especially deep learning (DL) models tailored specifically for image analysis and computer vision, into digital pathology workflow. These AI models are designed to assist pathologists with routine, time-consuming tasks, such as cell counting and screening large numbers of biopsies, as well as those with limited reproducibility, such as tumor grading and immunohistochemistry scoring.^3,4,6^ As such, AI represents a potential solution to address the shortage of pathologists by streamlining the diagnostic workflow.^7,8^

Artificial Neural Networks (ANNs) are softwares that mimic the connections in the human brain; put in series, they can be trained to specific tasks, such as recognizing a dog on a picture – or a melanoma on a slide. ANN can also be employed to produce new material, which can be categorized as Generative AI. In this burgeoning field, the development of large language models (LLMs) has marked a revolutionary breakthrough. According to the EU-U.S. terminology and taxonomy for AI, LLMs are “a class of language models that use deep-learning algorithms and are trained on extremely large textual datasets” and they have the unique capacity to decipher and generate human-like language with an unprecedented level of sophistication.^9^ These models are underpinned by multi-layered ANN and rely upon transformer architecture – a peculiar neural network structure introduced by Google researchers in 2017.^6,10–12^ In particular, LLMs are built by training with massive amounts of textual data (billions of words derived from the internet, including websites, academic books, scientific articles and websites), allowing them to recognize the patterns and structures of language by identifying relationships between words and phrases and, consequently, generate (by predicting linguistic patterns) natural human-like language text outputs that are contextually coherent to the input.^11–16^ The multi-stage training process involves varying degrees of human intervention and includes an unsupervised learning step (pre-training step) for language structure acquisition, followed by a supervised learning step (fine-tuning step) for task-specific optimization.^15,17^ The fine-tuning process is necessary to produce generative LLM chatbots with potential impactful applications, allowing this technology to draw attention in several fields, including the healthcare sector: ChatGPT, one of the most prominent and widely used LLM chatbots, is a prime example as it is currently generating great interest in the field of medical research.^11^ ChatGPT is a non-domain specific LLM developed in November 2022 by OpenAI that is capable of generating equal-to-human-level text in a chat-like setting. It is a specific application of the GPT-3 model – an LLM pre-trained on 175 billion parameters and on a large corpus of text data encompassing more than 400 billion words – that is fine-tuned over a transformer-based language model in the GPT-3.5 series (an improved version of GPT-3) via a process called reinforcement learning from human feedback (RLHF), a training modality that uses both human-assisted simulations and reward-based fine-tuning methods.^11,13–16,18–21^ Over the last year, LLMs, and in particular ChatGPT, have shown their great potential in physician-patient communication^22,23^, in medical administrative tasks such as writing discharge letters^23^, in assisting the medical educational process^14^, in medical research (e.g. academic literature analysis, clinical text data analysis, genetic and protein structure data analysis)^24–29^, in question answering regarding both medical student and specialist examinations^14,30,31^ and as a support tool for healthcare professionals in generating and interpreting clinical notes and diagnostic reports.^11,26,30^

The literature regarding the use of LLMs in pathology is limited.^32,33^ Despite this, various authors have highlighted the potential applications of LLMs in pathology for tasks such as classifying whole slides^34^, extracting information from pathology notes, creating more understandable pathology reports, summarizing patient data and research documents, generating differential diagnosis lists, and suggesting immunohistochemical stains and ancillary tests.^32,35^ Nevertheless, recent publications have started to document the application of ChatGPT in addressing pathology-related inquiries, engaging in clinical-pathological case analysis and assessing its potential role within molecular tumor board discussions.^33,36–40^ In the era of pioneering the application of AI in routine pathology, we decided to assess ChatGPT’s reliability in addressing pathology-related diagnostic questions by testing its performance on clinico-pathological scenarios, focusing our attention on its potential role as a supporting tool in the diagnostic process; secondly, since prompting strategies can significantly influence the quality and relevance of the generated outputs^41^, we also explored which prompting strategies would allow pathologists to converse most effectively with the LLM in order to receive adequate, coherent and evidence-based outputs.

## Material and methods

### Study design outlook

In order to assess the usefulness of ChatGPT as a support for pathological diagnosis, we created five clinico-pathological *scenarios* for ten pathological subspecialties (see next paragraph). Each clinical-pathological *scenario* was posed to ChatGPT using four different prompting strategies. Six pathologists then evaluated the *accuracy* and the *usefulness* of all ChatGPT outputs (*answers*). We also preregistered the study and performed a simulation-based calibration of the analytical pipeline prior to the pathologist evaluations. Study history, custom code and data can be found at the following repository: https://github.com/slrenne/PathGPT.

### Question design

A schematic representation of the prompt design is depicted in Figure 1. We designed prompts in a modular fashion, using different prompt patterns^41^; each prompt was framed as follows: (1) the *persona pattern* module contains a specific sentence that guides the LLM in determining the kind of output it should produce, focusing on details relevant to the output (“From now on act as a pathologist.”); (2) the *clinical scenario* module; (3) the *question type* module (multiple-choice or open-ended); (4) the r*eflection pattern* module contains a sentence that asks the LLM to explain the rationale behind a given output, uncovering underlying assumptions (“When you provide an answer, please explain the reasoning and assumptions behind it.”); (5) the *references type* module (with references or without references). We wrote the prompts as clinico-pathological scenarios, imagining a pathologist trying to use ChatGPT as an instrument to narrow down the differential diagnosis and solve the case. Scenarios were formulated from scratch by the authors in order to limit the possibility of their presence in the ChatGPT training dataset. We wrote 5 scenarios for 10 pathology subspecialties: 1) breast, 2) skin, 3) digestive system, 4) female genital, 5) hematolymphoid, 6) urinary and male genital, 7) endocrine and neuroendocrine, 8) central nervous system (CNS), 9) bone and soft tissue (BST) and 10) thoracic. Scenarios were written according to the most recent guidelines, reviewed by an expert pathologist in the subfield and corrected by a native English speaker (DNC). Each scenario was then prompted to ChatGTP using the following two types of question: 1) open-ended (OE) and 2) multiple-choice (MC). In the open ended questions, we elicited the answer by asking “Which are the most likely differential diagnoses?”. In the multiple-choice questions, we wrote five different differential diagnoses, imagining that the pathologist trying to narrow down the differentials might already have some. The position of the correct answer was randomized. Moreover, we followed the questions (OE or MC) with one of two options: 1) no further directives or requests (NR) or 2) asking for references (R) using the following statement: “All the statements of the explanation shall be followed by a reference (author, year) taken from medical and scientific literature. Please also list all the references at the end of the document (author, year, source).” Overall, we asked ChatGPT to provide scientific references two times for each scenario, once with the question in open-ended form and once with the question in multiple-choice form. All the questions were uploaded in the study repository.

**Figure 1.**
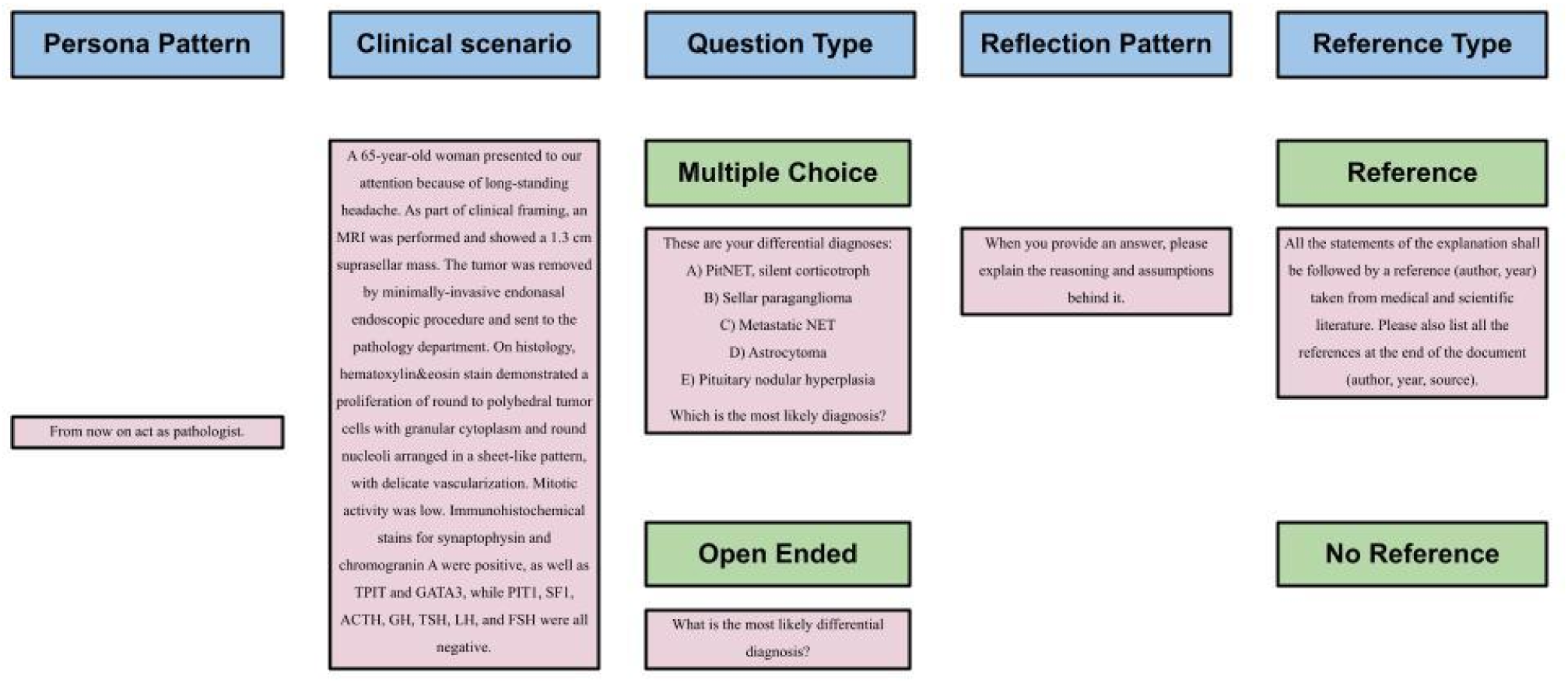
A schematic representation of the prompt design is shown. Clinico-pathological scenarios were designed in a modular fashion: the blue boxes denote the different modules used for each scenario, the green boxes indicate the four question modalities (MC-R, MC-NR, OE-R, OE-NR) while the remainings represent an example scenario. See also main text and online repository.

### Prompting

Each of the 200 questions (5 scenarios x 10 pathology subfields x 2 question-type prompts x 2 reference-type prompts) were submitted in a new chat session to reduce memory retention bias.^14^ The answers generated by ChatGPT were collected in a Google Form that recorded: (1) question ID; (2) scenario ID; (3) pathology subfield; (4) question type (MC or OE); (5) reference type (without scientific reference or with scientific reference); (6) question text; (7) answer text. All the answers can be found in the study repository.

### Evaluation

Six pathologists evaluated the answers generated by ChatGPT in a random order, assessing the usefulness of the answer in supporting the clinico-pathological diagnosis (useful, not useful) and counting the number of errors (i.e. the accuracy). The pathologists’ evaluations were collected in a Google Form that recorded: (1) question ID; (2) scenario ID; (3) pathology subfield; (4) question type; (5) reference type; (6) question and answer text; (7) usefulness (yes, no); (8) total number of errors (integer). Scientific references provided by ChatGPT were also manually verified.

### Statistics

We used a Bayesian Network to assess the knowledge of ChatGPT in Pathology; the graphical version is depicted in Figure 2. The latent knowledge (K) of ChatGPT in each subfield (F) of pathology was the estimand of interest. To probe this knowledge, clinical scenarios (S) were presented with distinct question types (QT) i.e. open ended/multiple choice and with or without reference inquiries (Ref). The responses (A) elicited from the model were evaluated by several pathologists (Pa), who assessed both the usefulness (U) and the number of errors (E) of each response. Additionally, the influence of the pathologist’s expertise in each specific field (F) was taken into account, as it may affect their evaluations.

**Figure 2.**
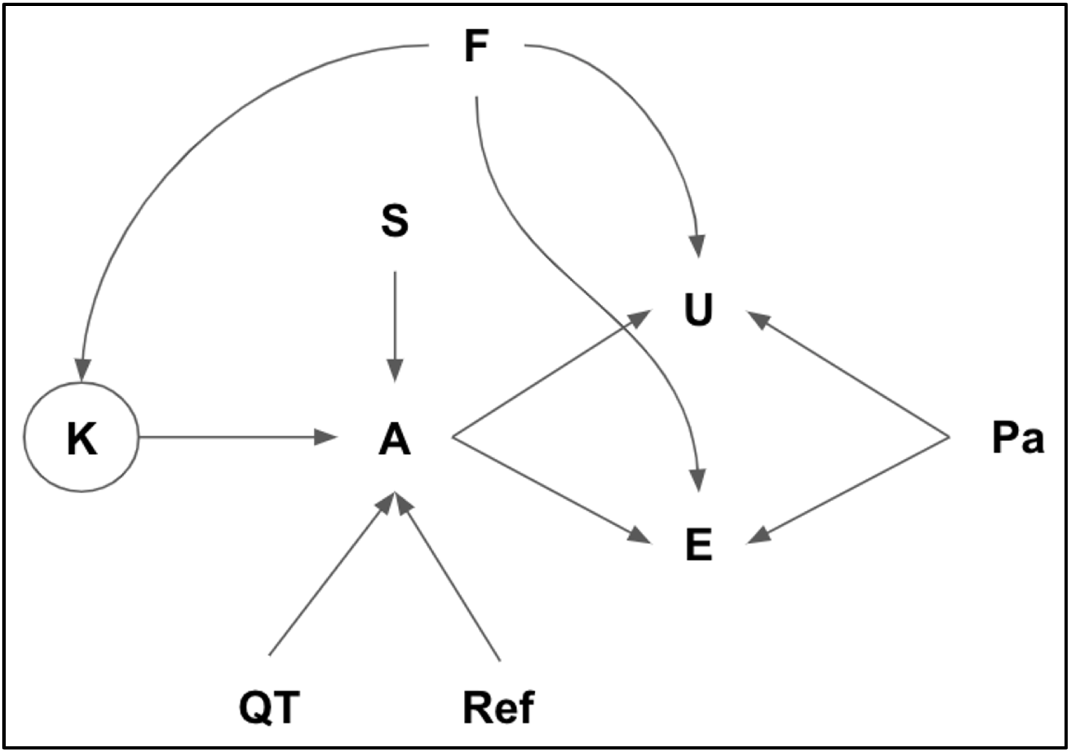
The relationship between the variables is shown. This directed acyclic graph (DAG) was then translated into a system of equations to determine the value of K. See text for further details.

### Multilevel hierarchical models

Usefulness (U) and errors (E) of the ChatGPT were modeled as:

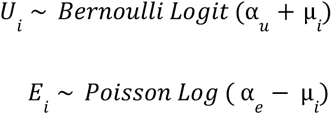

where µ_*i*_ is the effect of the answer (A) combined with the ‘pathologist effect’(ɛ):

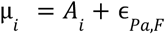

where ɛ represents the correlated variable of the Pathologist (Pa) and the subspecialty field (F). Finally the answer was modeled as follows:

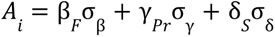

where β_*F*_ represents the latent knowledge for each subspecialty field (F), σ_β_ is the standard deviation of the knowledge in the various fields. Similarly, γ_*pr*_ estimates a parameter for each prompt combination (OE-R, OE-NR, MC-R, MC-NR), and σ_γ_ is the standard deviation of the prompt effect. Finally, δ_*S*_ estimates the effect of the different scenarios, and σ_δ_ is the standard deviation of this effect. Further model specification, priors and prior predictive simulations are available in the study repository.

### Model testing

Following the Bayesian workflow^42^, we generated simulated data according to the DAG depicted in Figure 2. We then used the simulated data to test the ability of the model to recover the parameters.^42^ Compatibility Intervals (CI) were computed as 89% Higher Posterior Density Region (HPDI). The model was run using R (Ver. 4.3.1) and Stan programming language (Ver. 2.26.1).^43,44^

### Qualitative analysis of best- and worst-case scenarios

The qualitative analysis of the three best- and three worst-case scenarios was conducted with the aim of testing whether ChatGPT’s performance depended on the presence (or absence) of clinical cases similar to those we formulated on the internet. For each of the diseases covered by the six scenarios, the analysis was initially conducted by Googling the text of the scenarios and, in the second instance, looking among the collection of published case-reports on the Pathology Outline website.^45^ The search conducted in this way allowed us to check if there were any published case reports on the internet, including patients with similarities to our clinico-pathological scenarios, both in terms of clinical presentation and histo-pathological, immunohistochemical and molecular biology findings.

## Results

### Gross outlook of responses

In the present study study, the 200 answers generated by ChatGPT were evaluated by 6 pathologists, yielding a total of 894 evaluations; of these, 450 (50.3%) were in open-ended form and 444 (49.7%) were in multiple-choice form, while (450) 50.3% did not require scientific references and 444 (49.7%) required them to be provided. ChatGPT provided useful answers in 556 cases (62.2%). 287 answers (32.1%) contained no errors, while 190 (21.3%) had one error, 133 (14.9%) had two errors, and 83 (9.3)% had three errors, with a maximum of 18 errors.

As regards scientific references, ChatGPT provided an average of 2.14 scientific references per response (range 1-5), for a total of 214 bibliographic references. Out of a them, 70.1% (150/214) were found to be true and correct, 12.1% (26/214) were found to be inaccurate (reporting errors regarding authors’ names, year of publication, journal of publication and/or number of pages given), while for 17.8% (38/214) of the references generated, we did not find a counterpart in the literature or they linked to completely different scientific work, although drafted and presented in a way that looked correct.

### Usefulness, accuracy latent knowledge, and prompting strategy

ChatGPT provided a useful answer in 61% of the cases (CI: 36% - 0.86%). The usefulness density was calculated with a posterior predictive simulation that excluded the pathologists’ effect (Figure 3A). Similarly, the expected number of errors was 1.86 (CI: 0.11 - 3.51), again without the pathologists’ effect. In this study, accuracy refers to ChatGPT’s ability to generate responses with fewer errors for a given question, indicating that the LLM produces more precise answers (Figure 3B). Latent knowledge across the ten fields showed minimal variation, with the lowest to highest order being urinary and male genital, female genital, hematolymphoid, CNS, BST, endocrine and neuroendocrine, breast, thoracic, digestive system, and skin pathology (Figure 3C). Finally, as regards prompting strategies, MC-NR proved to be the most effective;followed by MC-R, OE-NR and finally OE-R. Overall, ChatGPT performed slightly better in cases where we gave it answer options to choose from (i.e., differential diagnoses) and, for the same question type (OE or MC), in cases where we did not ask it to provide scientific references (Figure 3D).

**Figure 3.**
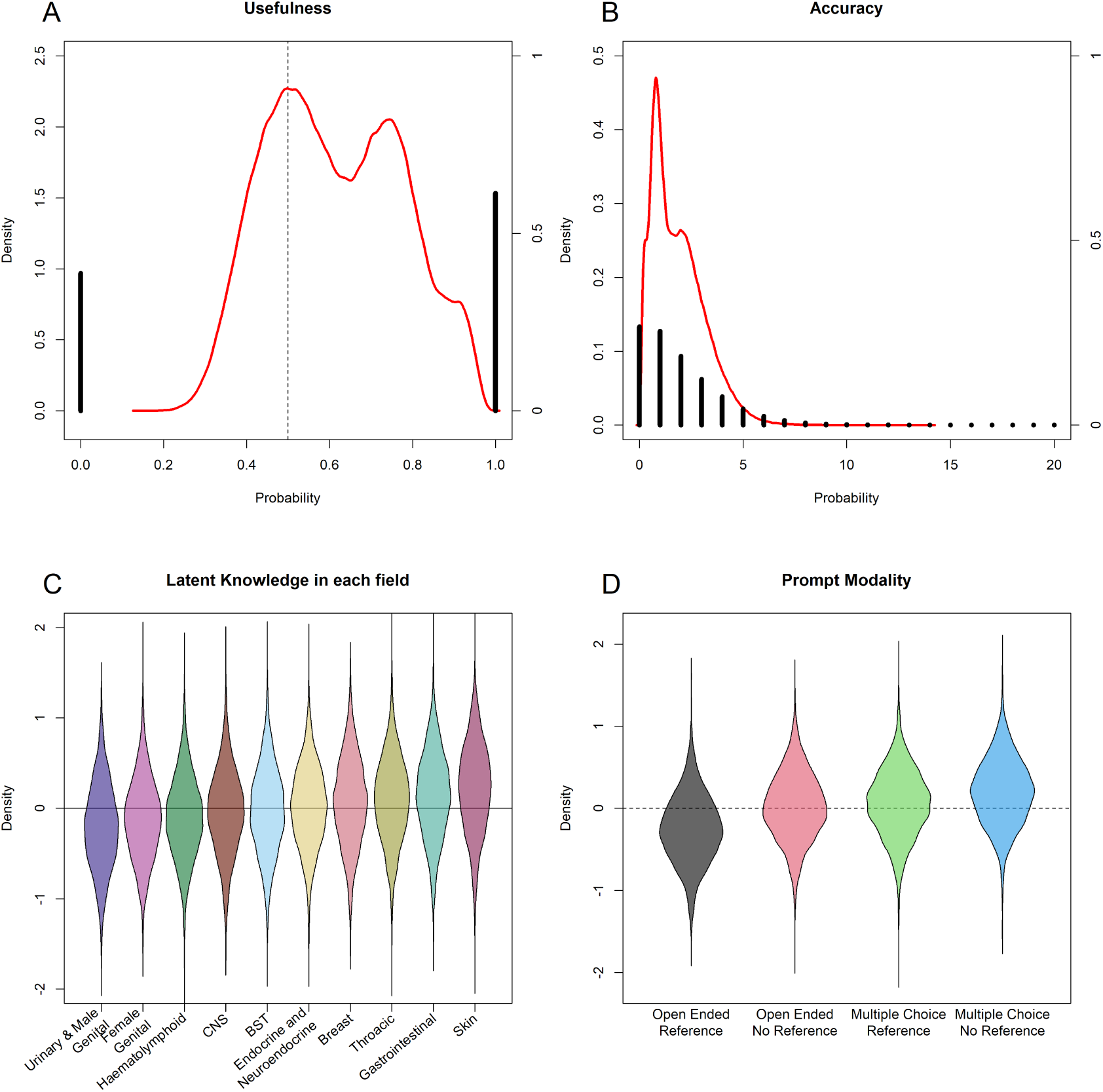
ChatGPT performances. (A) Usefulness of ChatGPT. Albeit considered useful, more often than not, the overall performance was not exceptional (red solid line is the posterior probability density; black histograms represent the proportion of cases useful (1) or non-useful (0), see right side of the figure for reference). B) Accuracy of ChatGPT. Similarly, the numbers of errors, although low, are substantial (red solid line is the posterior probability density; black histograms represent – for each number of errors – the proportion of cases, see right side of the figure for reference). (C) Knowledge of ChatGPT according to Fields. The model was designed to capture the latent knowledge of ChatGPT in each specific subfield. Although very small, the model was able to recover differences in the knowledge. (D) Performance of ChatGPT according to Prompt Types. Also prompting strategy did not produce striking changes in performance.

### Scenario had the greatest impact on ratings

Scenario variability had the greatest impact on ratings, followed by evaluator/subspecialty interactions, specific field knowledge and prompting strategy. The 50 clinico-pathological scenarios have a variable distribution in terms of probability density, which, in turn, accounts for the considerable impact they had on ChatGPT’s output assessment and, ultimately, on its performance (Figure 4). The three scenarios in which ChatGPT outperformed were the scenarios about pancreatic solid pseudopapillary neoplasm (scenario 34), sarcoidosis (scenario 36) and pilomatricoma (scenario 44); conversely, we recorded the worst ChatGPT performance in scenarios about histiocytic sarcoma (scenario 20), basosquamous carcinoma (scenario 41) and anaplastic pleomorphic xanthoastrocytoma (scenario 47) (Table 1).

**Figure 4.**
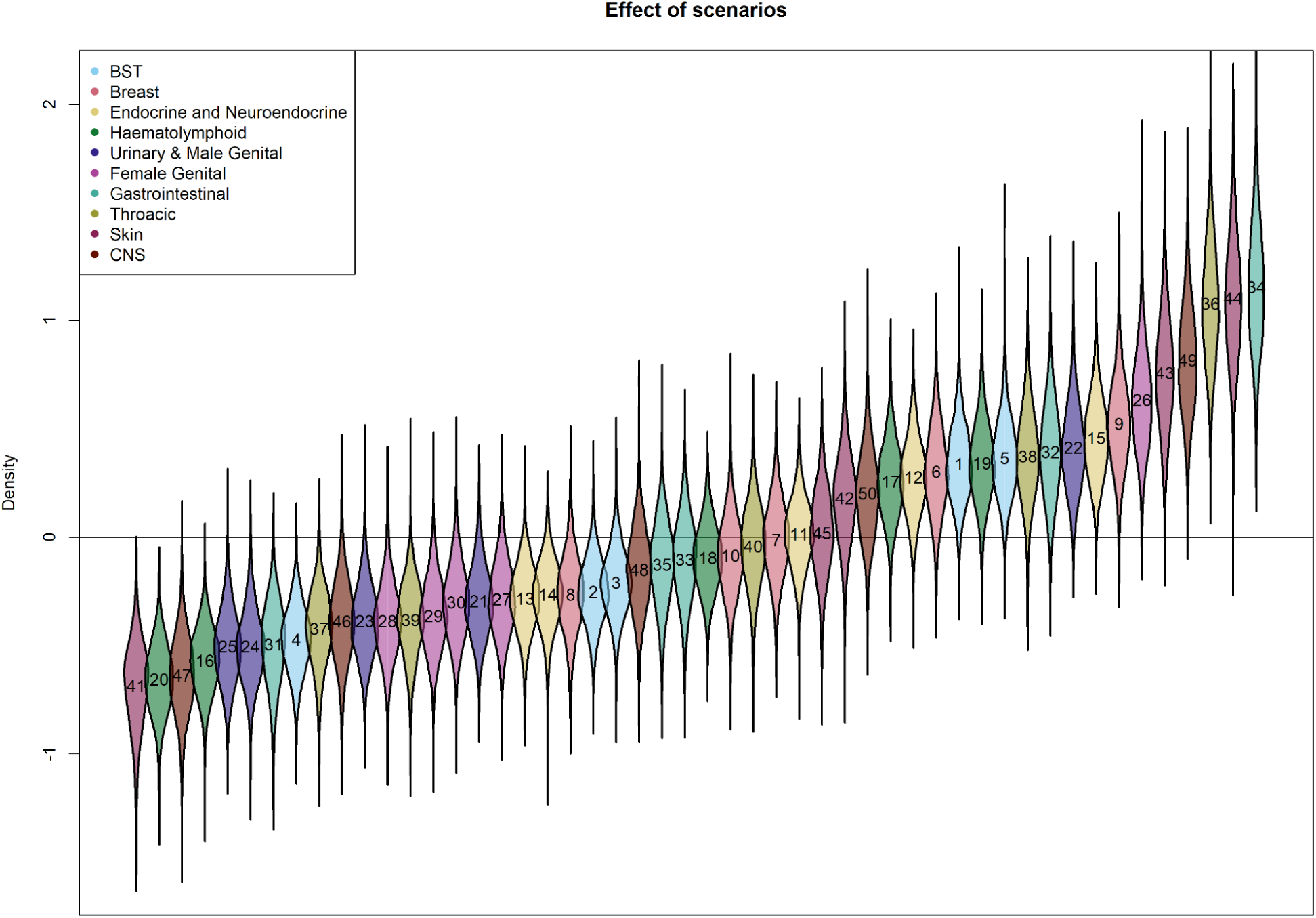
Effect of scenarios. Scenario had the most variability and the greatest impact on ratings.

**Table 1.**
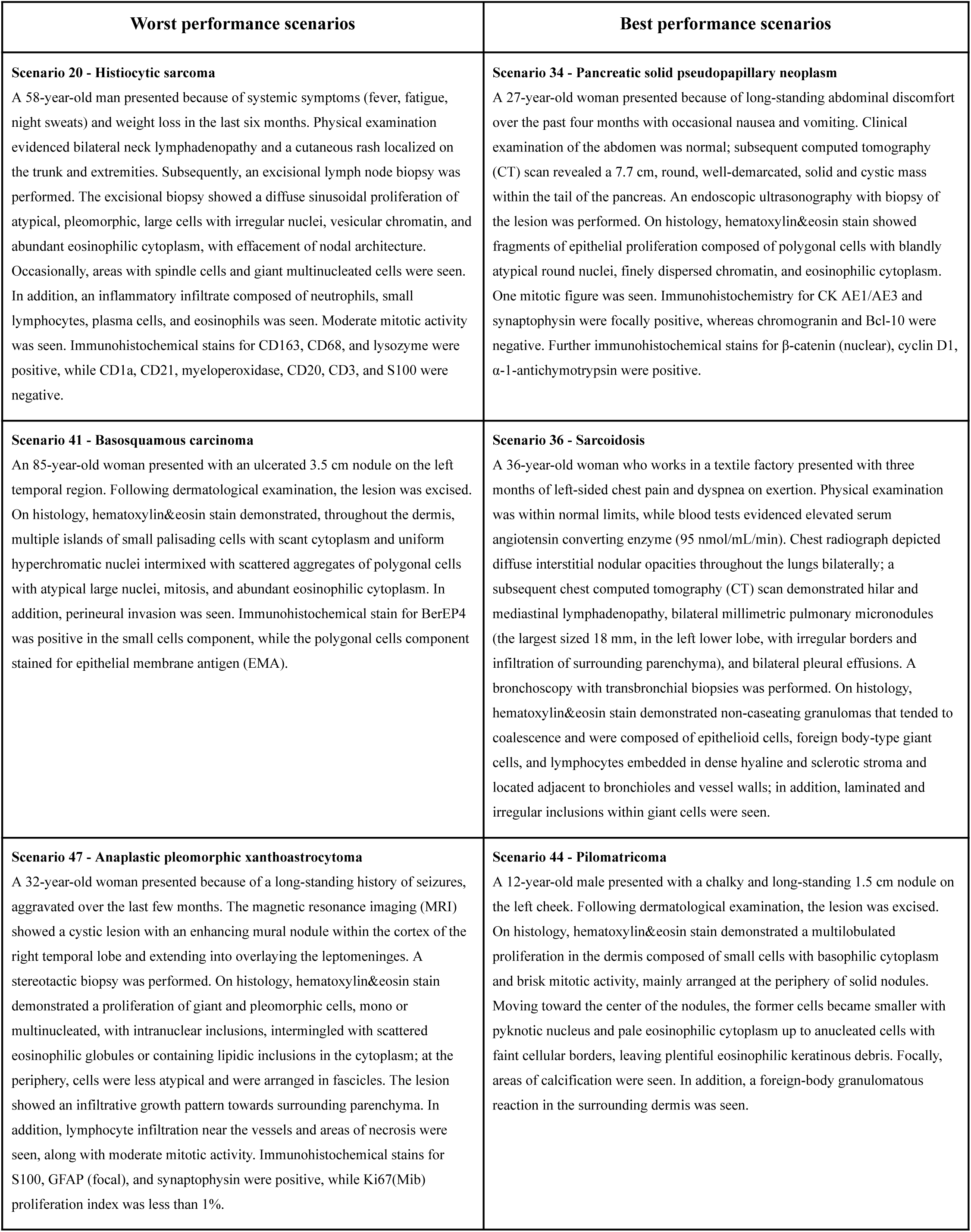
Best and worst performance scenarios.

| Worst performance scenarios | Best performance scenarios |
| --- | --- |
| **Scenario 20 - Histiocytic sarcoma**<br>A 58-year-old man presented because of systemic symptoms (fever, fatigue, night sweats) and weight loss in the last six months. Physical examination evidenced bilateral neck lymphadenopathy and a cutaneous rash localized on the trunk and extremities. Subsequently, an excisional lymph node biopsy was performed. The excisional biopsy showed a diffuse sinusoidal proliferation of atypical, pleomorphic, large cells with irregular nuclei, vesicular chromatin, and abundant eosinophilic cytoplasm, with effacement of nodal architecture. Occasionally, areas with spindle cells and giant multinucleated cells were seen. In addition, an inflammatory infiltrate composed of neutrophils, small lymphocytes, plasma cells, and eosinophils was seen. Moderate mitotic activity was seen. Immunohistochemical stains for CD163, CD68, and lysozyme were positive, while CD1a, CD21, myeloperoxidase, CD20, CD3, and S100 were negative. | **Scenario 34 - Pancreatic solid pseudopapillary neoplasm**<br>A 27-year-old woman presented because of long-standing abdominal discomfort over the past four months with occasional nausea and vomiting. Clinical examination of the abdomen was normal; subsequent computed tomography (CT) scan revealed a 7.7 cm, round, well-demarcated, solid and cystic mass within the tail of the pancreas. An endoscopic ultrasonography with biopsy of the lesion was performed. On histology, hematoxylin&eosin stain showed fragments of epithelial proliferation composed of polygonal cells with blandly atypical round nuclei, finely dispersed chromatin, and eosinophilic cytoplasm. One mitotic figure was seen. Immunohistochemistry for CK AE1/AE3 and synaptophysin were focally positive, whereas chromogranin and Bcl-10 were negative. Further immunohistochemical stains for β-catenin (nuclear), cyclin D1, α-1-antichymotrypsin were positive. |
| **Scenario 41 - Basosquamous carcinoma**<br>An 85-year-old woman presented with an ulcerated 3.5 cm nodule on the left temporal region. Following dermatological examination, the lesion was excised. On histology, hematoxylin&eosin stain demonstrated, throughout the dermis, multiple islands of small palisading cells with scant cytoplasm and uniform hyperchromatic nuclei intermixed with scattered aggregates of polygonal cells with atypical large nuclei, mitosis, and abundant eosinophilic cytoplasm. In addition, perineural invasion was seen. Immunohistochemical stain for BerEP4 was positive in the small cells component, while the polygonal cells component stained for epithelial membrane antigen (EMA). | **Scenario 36 - Sarcoidosis**<br>A 36-year-old woman who works in a textile factory presented with three months of left-sided chest pain and dyspnea on exertion. Physical examination was within normal limits, while blood tests evidenced elevated serum angiotensin converting enzyme (95 nmol/mL/min). Chest radiograph depicted diffuse interstitial nodular opacities throughout the lungs bilaterally; a subsequent chest computed tomography (CT) scan demonstrated hilar and mediastinal lymphadenopathy, bilateral millimetric pulmonary micronodules (the largest sized 18 mm, in the left lower lobe, with irregular borders and infiltration of surrounding parenchyma), and bilateral pleural effusions. A bronchoscopy with transbronchial biopsies was performed. On histology, hematoxylin&eosin stain demonstrated non-caseating granulomas that tended to coalescence and were composed of epithelioid cells, foreign body-type giant cells, and lymphocytes embedded in dense hyaline and sclerotic stroma and located adjacent to bronchioles and vessel walls; in addition, laminated and irregular inclusions within giant cells were seen. |
| **Scenario 47 - Anaplastic pleomorphic xanthoastrocytoma**<br>A 32-year-old woman presented because of a long-standing history of seizures, aggravated over the last few months. The magnetic resonance imaging (MRI) showed a cystic lesion with an enhancing mural nodule within the cortex of the right temporal lobe and extending into overlaying the leptomeninges. A stereotactic biopsy was performed. On histology, hematoxylin&eosin stain demonstrated a proliferation of giant and pleomorphic cells, mono or multinucleated, with intranuclear inclusions, intermingled with scattered eosinophilic globules or containing lipidic inclusions in the cytoplasm; at the periphery, cells were less atypical and were arranged in fascicles. The lesion showed an infiltrative growth pattern towards surrounding parenchyma. In addition, lymphocyte infiltration near the vessels and areas of necrosis were seen, along with moderate mitotic activity. Immunohistochemical stains for S100, GFAP (focal), and synaptophysin were positive, while Ki67(Mib) proliferation index was less than 1%. | **Scenario 44 - Pilomatricoma**<br>A 12-year-old male presented with a chalky and long-standing 1.5 cm nodule on the left cheek. Following dermatological examination, the lesion was excised. On histology, hematoxylin&eosin stain demonstrated a multilobulated proliferation in the dermis composed of small cells with basophilic cytoplasm and brisk mitotic activity, mainly arranged at the periphery of solid nodules. Moving toward the center of the nodules, the former cells became smaller with pyknotic nucleus and pale eosinophilic cytoplasm up to anucleated cells with faint cellular borders, leaving plentiful eosinophilic keratinous debris. Focally, areas of calcification were seen. In addition, a foreign-body granulomatous reaction in the surrounding dermis was seen. |

Considering the impact that scenario variability had on the evaluation of ChatGPT outputs, we decided to conduct a text quality analysis of the three scenarios where ChatGPT showed superior results and of the three scenarios in which it performed worst to see if similar scenarios had already been reported in the published literature, so as to explain, at least in part, its varying levels of accuracy. The research did not evidence similar published cases to any of these fictional scenarios.

## Discussion

Overall, although ChatGPT outputs were evaluated as useful in supporting a pathological diagnosis in 61% of cases, the absolute number of errors remains high in about half of the outputs, with only one-third of the outputs free of errors. Hence, caution is necessary when considering it as a diagnostic support tool; therefore, the free version of ChatGPT is not yet adequate for systematic use in everyday diagnostic practice. In addition, the absolute number of errors is on average higher for more experienced assessors – that is, those most likely to face routine diagnostics – and this further negatively affects the potential usefulness of ChatGPT in routine diagnostics.

In the present study, we have extensively questioned ChaGPT on clinico-pathological scenarios to assess its potential usefulness in supporting pathologists in their daily diagnostic workload. What emerges is that what is asked (i.e. the different pathological conditions in each scenario) had the greatest impact, whereas specific field knowledge and prompting strategy are of little influence. The three scenarios in which ChatGPT outperformed were the scenarios about pancreatic solid pseudopapillary neoplasm (GI), sarcoidosis (Thoracic) and pilomatricoma (Skin); conversely, we recorded the worst ChatGPT performance in scenarios about histiocytic sarcoma (Hemolymph), basosquamous carcinoma (Skin) and anaplastic pleomorphic xanthoastrocytoma (CNS). These results led us to make some considerations: most importantly, that it is surprising to see that ChatGPT performed better in scenarios reporting diseases with relatively low prevalence given ChatGPT’s mode of (pre)-training on a vast amount of text data from the internet. However, in our qualitative analysis, we did not find evidence of similar published cases.

As previously mentioned, prompting strategy had the lowest impact on output rating. The fact that ChatGPT performed slightly better in clinico-pathological questions with five differential diagnoses to choose from could suggest that doing so narrows the range of diagnostic hypotheses to which ChatGPT must associate the clinico-pathological presentation of the scenario in order to provide the statistically most probable diagnosis. Indeed, in pathology it is not uncommon for different pathologies to present with similar clinical manifestations and overlapping histological features. This suggests that, in the future, the most appropriate potential use for an LLM in diagnostic pathology might be that of a “digital assistant” who provides a second opinion on real-life clinico-pathological cases of particular diagnostic complexity – on the basis of some diagnostic hypothesis that implies structured diagnostic reasoning by the requesting pathologist – rather than that of an "intelligent oracle" to whom one can turn to solve a challenging diagnostic question in the absence of clinical-diagnostic reasoning. Finally, the fact that prompting strategies largely overlap in terms of output rating could suggest that it is not the way of posing the clinical-pathological scenario to ChatGPT (i.e. in an open-ended form or providing possible differential diagnoses) that influences the quality of the output, but rather what pathological topic the LLM is asked about and its level of training on that specific topic.

With regard to bibliographic scientific references, our results pointed to a substantial lack of accuracy on the part of ChatGPT, further confirming that extreme care must be taken in its use not only as a diagnostic support tool but in all academic spheres and that, in any case, any output should at least be double-checked. In addition, these results were compared with the published scientific literature and are in line with what other authors have reported, who describe a consistent lack of thoroughness in the generation of references.^46–49^

In recent years, interest in LLMs has grown exponentially, and this phenomenon has had repercussions in the biomedical field as well. Prior to the official release of ChatGPT, several authors tested the performance of different LLMs (GPT-3.5 (Codex and InstructGPT), BERT, RoBERTa, ALBERT, PaLM, PubMedGPT, etc.) against multiple-choice medical licensing exam questions, reaching varying levels of accuracy, sometimes achieving human-level performance.^50–52^ Since the release date of ChatGPT, multiple papers have been published investigating performance and accuracy in answering questions belonging to a wide variety of medical subspecialties, often comparing AI performance to that obtained by humans on the same question sets.^14,37,49,53–60^ Most of these studies were conducted using standardized, pre-formulated questions picked from board medical licensing exam question pools (i.e. United Medical Licensing Exam (USMLE), Ophthalmic Knowledge Assessment Program (OKAP) exam, American Heart Association (AHA), Chinese National Medical Licensing Examination)^14,49,56,60^, written based on university textbooks^59^, reproduced from published case reports^54^ or retrieved from social media.^57^ These studies typically discarded or re-described questions containing photographs, graphs, figures and tables and subsequently posed them to ChatGPT in open-ended or multiple-choice form. Albeit with some exceptions^49,54,60^, most of the works available in literature found an overall accuracy around or above 50% in the majority of the experiments – equaling, in some cases, human performance with respect to recall and knowledge – and that LLMs provided insightful explanations to support its answer choices, especially in scenario-based questions.^14,55,56^ Interestingly, Ming Liang Oon et al. are among the first to have evaluated the utility of ChatGPT4 in routine diagnostics by submitting 10 challenging virtual slides to nine junior pathologists and trainees (with fewer than 10 years of experience) and comparing their performance with and without ChatGPT assistance.^37^ Similar to our findings, they reported that ChatGPT’s diagnostic accuracy was inferior to that of pathologists and that consultation with ChatGPT yielded only marginal benefit. They also reported that ChatGPT outputs were heavily influenced by the prompting; however, it is worth noting that their use of the term is wider and ecompasses the scenario effect as well as the prompting strategy. We can state that our results, in terms of accuracy, are in line with what has been reported in the literature; in fact, ChatGPT generated outputs rated as error-free or with one error in 53.2% of the evaluations (32% and 21.2% respectively).^1^

Our study presents few limitations, namely that, at the time we conceptualized and began the study, ChatGPT did not yet have the ability to analyze images. For this reason, we clearly spelled out all necessary elements to render a pathological diagnosis (as reported in the reference texts for diagnostic practice), since, after all, ChatGPT is designed and trained to handle texts. Similarly, we used the free version of ChatGPT, thus, not the most up-to-date model (its training dataset comprised materials generated up until 2021); this was the only option because it was the only available version when the study began. Finally, it is necessary to consider that using human’s assessments as a metric for accuracy can be biased. Hence, we involved multiple evaluators along with a multilevel hierarchical model that allowed us to make robust inferences on accuracy and usefulness and to obtain the estimate of accuracy and usefulness free of human ( pathologist) effect.

This study underscores the limitations of ChatGPT’s potential as a diagnostic support tool. While ChatGPT demonstrated utility in certain scenarios, the variation in its performance underlines the need for cautious application, highlighting the importance of further development and refinement of LLM tools before their introduction into routine practice, especially in their ability to adapt to the dynamic nature of medical knowledge and to interact more effectively with physicians. The study also suggests that, in the future, LLMs could be involved in the diagnostic process as adjunct assistants in complex and specific contexts, particularly where pathologists require a secondary opinion or a quick reference; in this regard, creating specialized LLMs through a subspecialty-specific training process could lead to improved performance and accuracy to the extent that it could significantly improve usefulness in routine pathological diagnostic activity. However, the variability in error rates indicates that ChatGPT is not yet a substitute for human judgment in pathology. In addition, given the variability in error rates and the inaccuracy in providing scientific reference, utmost caution is also necessary when considering it as a self-learning or self-assessing tool.

In conclusion, this study has critically evaluated the usefulness of ChatGPT in the surgical pathology diagnostic process, addressing two primary research questions and offering insights into the future potential role and limitations of using LLMs in the pathologic field. Our findings indicate that discretion is necessary when considering ChatGPT as a diagnostic support tool and that it is not yet adequate for use in everyday diagnostic practice. Indeed, its effectiveness is influenced by the variability of clinical scenarios and, while showing competence in certain clinico-pathological scenarios, the overall error rate suggests a need for cautious application. In addition, our findings about the effect of the prompt strategy on ChatGPT’s performance underscores what might be the future modalities by which a pathologist could interrogate LLMs. Moreover, the lack of thoroughness in providing scientific references together with the reported error rates suggest that caution should be employed in using ChatGPT even as a self-learning or self-assessing tool. Finally, this study confirms that ChatGPT can achieve a level of performance and accuracy similar to that observed in other scientific works. Therefore, all the results we have obtained confirm that it is essential to recognize the irreplaceable role of human experts who integrate images, clinical data and extensive experience, and who can still surpass Large Language Models in the intricate task of histopathological diagnosis.

## Data Availability

All data produced are available online at https://github.com/slrenne/PathGPT

## Footnotes

1 Aiming to test the potential of ChatGPT in routine diagnostic practice, we believe that quantifying the usefulness too as a performance measurement is appropriate for our study. Also, we strongly believe that any LLM, as of now, should be utilized as a mere assistant to any medical professional. However, as an orthodox trend, we considered the term accuracy is inversely proportional to number of errors.

## References

1. Zehra, T., Parwani, A., Abdul-Ghafar, J. & Ahmad, Z. A suggested way forward for adoption of AI-Enabled digital pathology in low resource organizations in the developing world. Diagn. Pathol. 18, 68 (2023).

2. Sung, H., et al. Global Cancer Statistics 2020: GLOBOCAN Estimates of Incidence and Mortality Worldwide for 36 Cancers in 185 Countries. CA: Cancer J. Clin. 71, 209–249 (2021).

3. Song, A. H. et al. Artificial intelligence for digital and computational pathology. Nat Rev Bioeng 1, 930–949 (2023).

4. Kim, I., Kang, K., Song, Y. & Kim, T.-J. Application of Artificial Intelligence in Pathology: Trends and Challenges. Diagnostics (Basel) 12, 2794 (2022).

5. Jahn, S. W., Plass, M. & Moinfar, F. Digital Pathology: Advantages, Limitations and Emerging Perspectives. J. Clin. Med. 9, 3697 (2020).

6. Waqas, A. et al. Revolutionizing Digital Pathology With the Power of Generative Artificial Intelligence and Foundation Models. Lab Invest 103, 100255 (2023).

7. Asif, A. et al. Unleashing the potential of AI for pathology: challenges and recommendations. J. Pathol. 260, 564–577 (2023).

8. Jurmeister, P., Müller, K.-R. & Klauschen, F. Artificial intelligence: a solution for the lack of pathologists? Pathologe 43, 218–221 (2022).

9. EU-U.S. Terminology and Taxonomy for Artificial Intelligence (first edition). https://digital-strategy.ec.europa.eu/en/library/eu-us-terminology-and-taxonomy-artificial-intelligence (2023).

10. Qwiklabs-Courses. Introduction to Generative AI. Google Cloud Skills Boost https://www.cloudskillsboost.google/course_sessions/4571341/video/434966 (2023).

11. Thirunavukarasu, A. J. et al. Large language models in medicine. Nat. Med. 29, 1930–1940 (2023).

12. Vaswani, A., et al. Attention Is All You Need. Preprint at 10.48550/arXiv.1706.03762 (2023).

13. Brown, T. B., et al. Language Models are Few-Shot Learners. Preprint at 10.48550/arXiv.2005.14165 (2020).

14. Kung, T. H. et al. Performance of ChatGPT on USMLE: Potential for AI-assisted medical education using large language models. PLOS Digit. Health 2, e0000198 (2023).

15. Radford, A. et al. Language Models are Unsupervised Multitask Learners. OpenAI blog 1, 9 (2019).

16. OpenAI. OpenAI Platform. https://platform.openai.com https://platform.openai.com/docs/models/overview.

17. Devlin, J., Chang, M.-W., Lee, K. & Toutanova, K. BERT: Pre-training of Deep Bidirectional Transformers for Language Understanding. Preprint at 10.48550/arXiv.1810.04805 (2019).

18. OpenAI. Introducing ChatGPT. https://openai.com/ https://openai.com/blog/chatgpt (2022).

19. OpenAI. OpenAI Platform. https://platform.openai.com https://platform.openai.com/docs/guides/gpt.

20. Christiano, P. F., et al. Deep Reinforcement Learning from Human Preferences. Preprint at 10.48550/arXiv.1706.03741 (2023).

21. Stiennon, N. et al. Learning to summarize with human feedback. in Advances in Neural Information Processing Systems vol. 33 3008–3021 (Curran Associates, Inc., 2020).

22. Ayers, J. W. et al. Comparing Physician and Artificial Intelligence Chatbot Responses to Patient Questions Posted to a Public Social Media Forum. JAMA Intern. Med. 183, 589–596 (2023).

23. Ali, S. R., Dobbs, T. D., Hutchings, H. A. & Whitaker, I. S. Using ChatGPT to write patient clinic letters. *Lancet Digit*. Health 5, e179–e181 (2023).

24. Yang, X. et al. A large language model for electronic health records. *npj Digit*. Med. 5, 194 (2022).

25. Agrawal, M., Hegselmann, S., Lang, H., Kim, Y. & Sontag, D. Large Language Models are Few-Shot Clinical Information Extractors. Preprint at 10.48550/arXiv.2205.12689 (2022).

26. Huang, K., Altosaar, J. & Ranganath, R. ClinicalBERT: Modeling Clinical Notes and Predicting Hospital Readmission. Preprint at 10.48550/arXiv.1904.05342 (2020).

27. Madani, A. et al. Large language models generate functional protein sequences across diverse families. Nat. Biotechnol. 41, 1099–1106 (2023).

28. Mai, D. H. A., Nguyen, L. T. & Lee, E. Y. TSSNote-CyaPromBERT: Development of an integrated platform for highly accurate promoter prediction and visualization of Synechococcus sp. and Synechocystis sp. through a state-of-the-art natural language processing model BERT. Front. Genet. 13, 1067562 (2022).

29. Jumper, J. et al. Highly accurate protein structure prediction with AlphaFold. Nature 596, 583–589 (2021).

30. Sallam, M. ChatGPT Utility in Healthcare Education, Research, and Practice: Systematic Review on the Promising Perspectives and Valid Concerns. Healthcare (Basel) 11, 887 (2023).

31. Nori, H., King, N., McKinney, S. M., Carignan, D. & Horvitz, E. Capabilities of GPT-4 on Medical Challenge Problems. Preprint at 10.48550/arXiv.2303.13375 (2023).

32. Malik, S. & Zaheer, S. ChatGPT as an aid for pathological diagnosis of cancer. Pathol. Res. Pract. 253, 154989 (2024).

33. Schukow, C. et al. Application of ChatGPT in Routine Diagnostic Pathology: Promises, Pitfalls, and Potential Future Directions. Adv. Anat. Pathol. 31, 15–21 (2024).

34. Pisula, J. I. & Bozek, K. Language models are good pathologists: using attention-based sequence reduction and text-pre-trained transformers for efficient WSI classification. Preprint at 10.48550/arXiv.2211.07384 (2022).

35. Hart, S. N. et al. Organizational preparedness for the use of large language models in pathology informatics. J Pathol Inform 14, 100338 (2023).

36. Cazzato, G. et al. Chat GPT in Diagnostic Human Pathology: Will It Be Useful to Pathologists? A Preliminary Review with ‘Query Session’ and Future Perspectives. AI 4, 1010–1022 (2023).

37. Oon, M. L., Syn, N. L., Tan, C. L., Tan, K.-B. & Ng, S.-B. Bridging bytes and biopsies: A comparative analysis of ChatGPT and histopathologists in pathology diagnosis and collaborative potential. Histopathology 84, 601–613 (2024).

38. Sinha, R. K., Deb Roy, A., Kumar, N. & Mondal, H. Applicability of ChatGPT in Assisting to Solve Higher Order Problems in Pathology. Cureus 15, e35237 (2023).

39. Vaidyanathaiyer, R. et al. Navigating the path to precision: ChatGPT as a tool in pathology. Pathol. Res. Pract. 254, 155141 (2024).

40. Sorin, V. et al. Large language model (ChatGPT) as a support tool for breast tumor board. NPJ Breast Cancer 9, 44 (2023).

41. White, J., et al. A Prompt Pattern Catalog to Enhance Prompt Engineering with ChatGPT. Preprint at 10.48550/arXiv.2302.11382 (2023).

42. Gelman, A., et al. Bayesian Workflow. Preprint at 10.48550/arXiv.2011.01808 (2020).

43. R Core Team. R: A Language and Environment for Statistical Computing. R Foundation for Statistical Computing (2023).

44. Stan Development Team. Stan Modeling Language Users Guide and Reference Manual. (2023).

45. Pathology Outlines - PathologyOutlines.com. https://www.pathologyoutlines.com/.

46. Manohar, N. & Prasad, S. S. Use of ChatGPT in Academic Publishing: A Rare Case of Seronegative Systemic Lupus Erythematosus in a Patient With HIV Infection. Cureus 15, e34616 (2023).

47. Akhter, H. M. & Cooper, J. S. Acute Pulmonary Edema After Hyperbaric Oxygen Treatment: A Case Report Written With ChatGPT Assistance. Cureus 15, e34752 (2023).

48. Benoit, J. R. A. ChatGPT for Clinical Vignette Generation, Revision, and Evaluation. Preprint at 10.1101/2023.02.04.23285478 (2023).

49. Fijačko, N., Gosak, L., Štiglic, G., Picard, C. T. & John Douma, M. Can ChatGPT pass the life support exams without entering the American heart association course? Resuscitation 185, 109732 (2023).

50. Liévin, V., Hother, C. E., Motzfeldt, A. G. & Winther, O. Can large language models reason about medical questions? Preprint at 10.48550/arXiv.2207.08143 (2023).

51. Jin, D. et al. What Disease Does This Patient Have? A Large-Scale Open Domain Question Answering Dataset from Medical Exams. Appl. Sci. 11, 6421 (2021).

52. Ha, L. A. & Yaneva, V. Automatic Question Answering for Medical MCQs: Can It Go Further than Information Retrieval? in Proceedings - Natural Language Processing in a Deep Learning World 418–422 (Incoma Ltd., Shoumen, Bulgaria, 2019).doi:10.26615/978-954-452-056-4_049.

53. Mbakwe, A. B., Lourentzou, I., Celi, L. A., Mechanic, O. J. & Dagan, A. ChatGPT passing USMLE shines aspotlight on the flaws of medical education. *PLOS Digit*. Health 2, e0000205 (2023).

54. Huh, S. Are ChatGPT’s knowledge and interpretation ability comparable to those of medical students in Korea for taking a parasitology examination?: a descriptive study. J. Educ. Eval. Health Prof. 20, 1 (2023).

55. Gilson, A., et al. How Does ChatGPT Perform on the United States Medical Licensing Examination? The Implications of Large Language Models for Medical Education and Knowledge Assessment. JMIR Med. Educ. 9, e45312 (2023).

56. Antaki, F., Touma, S., Milad, D., El-Khoury, J. & Duval, R. Evaluating the Performance of ChatGPT in Ophthalmology: An Analysis of Its Successes and Shortcomings. Ophthalmol. Sci. 3, 100324 (2023).

57. Duong, D. & Solomon, B. D. Analysis of large-language model versus human performance for genetics questions. Eur. J. Hum. Genet. (2023) doi:10.1038/s41431-023-01396-8.

58. Yeo, Y. H. et al. Assessing the performance of ChatGPT in answering questions regarding cirrhosis and hepatocellular carcinoma. Clin Mol Hepatol 29, 721–732 (2023).

59. Nisar, S. & Aslam, M. S. Is ChatGPT a Good Tool for T&CM Students in Studying Pharmacology? SSRN Scholarly Paper at 10.2139/ssrn.4324310 (2023).

60. Wang, X. et al. ChatGPT Performs on the Chinese National Medical Licensing Examination. J. Med. Syst. 47, 86 (2023).

